# Hypertension Trends and Disparities over Twelve Years in a Large Health System: Leveraging the Electronic Health Records

**DOI:** 10.1101/2023.08.24.23294518

**Authors:** John E. Brush, Yuan Lu, Yuntian Liu, Jordan R. Asher, Shu-Xia Li, Mitsuaki Sawano, Patrick Young, Wade L. Schulz, Mark Anderson, John S. Burrows, Harlan M. Krumholz

## Abstract

**Background:** The digital transformation of medical data enables health systems to leverage real-world data (RWD) from electronic health records (EHR) to gain actionable insights for improving hypertension care.

**Methods:** We performed a serial cross-sectional analysis of outpatients of a large regional health system from 2010 to 2021. Hypertension was defined by systolic blood pressure (SBP) ≥ 140 mmHg or diastolic blood pressure (DBP) ≥ 90 mmHg) or recorded treatment with anti-hypertension medications. We evaluated four methods of using blood pressure measurements in the EHR to define hypertension.

The primary outcomes were age-adjusted prevalence rates and age-adjusted control rates. Secondary outcomes were age-adjusted mean SBP and DBP and age-adjusted proportion of patients with a searchable diagnosis code of hypertension in the EHR.

**Results:** Hypertension prevalence varied depending on the definition used, ranging from 36.5% to 50.9% initially and increasing over time by approximately 5%, regardless of the definition used. Control rates ranged from 61.2% to 71.3% initially, rose during 2018-2019 and fell during 2020-2021. The proportion of patients with a hypertension diagnosis ranged from 45.5% to 60.2% initially and improved during the study period. Non-Hispanic Black patients represented 25% of our regional population and consistently had higher prevalence rates, higher mean SBP and DBP and lower control rates compared with other racial and ethnic groups.

**Conclusion:** In a large regional health system, we leveraged the EHR to provide real-world insights. The findings largely reflected national trends but showed distinctive regional demographics and findings. The findings have provided opportunities for improvement, with prevalence increasing, a quarter of the patients not controlled, and marked disparities. This approach could be emulated by regional health systems seeking to improve hypertension care.

**Key Points:** **Question:** Can a large regional health system leverage the electronic health record to analyze hypertension trends and disparities to drive improvement?

**Findings:** We analyzed 1,376,325 patients over 12 years and found that age-adjusted hypertension prevalence increased by approximately 5%. Age-adjusted hypertension control rates were in the 70% range and remained stable. Non-Hispanic Black patients represented 25% of our specific regional population and had 12-14% higher hypertension prevalence rates, higher mean age-adjusted systolic and diastolic blood pressure, and lower hypertension control rates compared with other racial groups.

**Meaning:** Real world data can provide actionable insights about hypertension and disparities in a specific region that could inform regional system strategies and initiatives for improvement.

## Introduction

Hypertension is a persistent and challenging health problem in the United States, estimated to affect almost half of adult Americans.^1^ Of patients with hypertension, approximately half have uncontrolled blood pressure (BP),^2^ which disproportionately affects African Americans.^2–4^ Improving the detection and treatment hypertension is a national priority and offers an important opportunity to modify the risk for cardiovascular disease and stroke and to address a major racial disparity.^5^

A 2014 advisory from the American College of Cardiology, American Heart Association and the US Centers for Disease Control and Prevention called for health system approaches for addressing hypertension.^6^ System-wide strategies, along with improvements at the clinician and patient level, could help optimize BP control and effectively modify the risk associated with hypertension.^6.7^ To help design health system strategies, more information is needed to characterize hypertension trends and identify opportunities for improvement at the regional health system level, ideally utilizing readily available real-world evidence from the electronic health record (EHR) systems.

There are opportunities to broaden the ways a regional health system might use EHR data to investigate hypertension. Converting EHR data into a common data model can provide a more agile analytical platform, which could enable a health system to take advantage of their long-term data, explore various analytical methods, and evaluate trends and demographics that may be unique to a health system’s region. Analysis of data at the regional level could enable customized regional initiatives to improve hypertension care.

Accordingly, we leveraged the EHR from a large health system caring for a diverse population to evaluate performance in the care of patients with hypertension.^8^ EHR data over 12 years were transformed into a harmonized and validated data set that has enabled a serial cross-sectional analysis of hypertension in a large regional population.^9^ We specifically sought to identify trends using various operational hypertension definitions to investigate disparities and opportunities for improvement.

## Methods

### Data Source

This retrospective observational study was performed at a large non-profit integrated healthcare system in Virginia and Northeastern North Carolina. The system began using a centralized EHR system designed by the Epic Corporation in 2007. In 2021, the health system began extracting key clinical data from its EHR and transforming the data into a harmonized data platform using the Observational Medical Outcomes Partnership (OMOP) common data model version 5.3.^9^

### Study Population

The overall study population consisted of all adult patients (≥18 years old) who had at least 1 outpatient blood pressure reading recorded between January 1st, 2010, and December 31st, 2021. Emulating the NHANES study,^2^ we conducted a serial cross-sectional study by independently analyzing the data in 2-year cycles from 2010-2011 to 2020-2021. In contrast to the defined clinical protocols of NHANES, our real-world data approach required defining computational protocols to analyze recorded BP readings in the EHR to create operational definitions of hypertension.

BP readings were first analyzed at the visit level. If more than one BP reading was taken during a visit, the first reading was disregarded and the mean of the remaining BP readings from the visit defined the visit BP measurement. If multiple outpatient visits occurred for a patient on a specific date, we calculated the mean of the visit BP measurements and used the mean as the visit BP measurement for that date.

### Definition of Hypertension

In each 2-year cycle, a patient was classified as having hypertension according to the definitions listed below using either an elevated visit BP measurement, or use of at least one first line anti-hypertension medication recorded during the 2-year cycle, like NHANES.^2^ BP elevation was defined as a systolic blood pressure (SBP) ≥ 140 mm Hg, or diastolic blood pressure (DBP) ≥ 90 mm Hg. First-line anti-hypertension medications were defined according to the 2017 AHA/ACC hypertension guideline^10^ and are listed with their associated OMOP concept identification numbers in eTable 1.

We evaluated four operational hypertension definitions using four different methods for incorporating recorded BP measurement into the definition: 1) using at least one elevated visit BP measurement during a 2-year cycle, 2) using the first visit BP measurement during a 2-year cycle, 3) using a single randomly chosen visit BP measurement during a 2-year cycle, or 4) using at least two elevated visit BP measurements during a 2-year cycle. Our rationale for using these different operational hypertension definitions was to provide definitions that are comparable with other studies and guideline recommendations. For example, our third definition would be comparable to the NHANES study,^2^ and our fourth definition would be comparable to the generally accepted guideline definition.^10^

### Definitions of Outcomes

Primary outcomes were age-adjusted hypertension prevalence and age-adjusted rates of BP control during each 2-year cycle. BP control was defined at the patient level as a mean SBP < 140 mmHg and mean DBP < 90 mmHg during a 2-year cycle. To adjust for differences in age distribution among the 2-year study cycles, direct standardization was employed using two different standards. For age-adjustment of hypertension prevalence and average BP, the standard was the average age distribution of all adults across all study cycles, while for age-adjustment of other outcome measures, the standard was average age distribution of adults with hypertension across all study cycles. The age categories used for standardization were: 18-44 years, 45-64 years, 65-74 years, and 75 years or older. The proportions used for age-adjustment of all patients and of hypertension patients for each operational hypertension definition are listed in eTable 2.

Secondary outcome measures were the age-adjusted mean SBP and DBP for all patients and for hypertensive patients, and the age-adjusted proportions of patients who were labelled with a searchable diagnosis code of hypertension in the EHR during a 2-year cycle. The coded diagnoses of hypertension and the corresponding OMOP concept identification numbers listed in eTable 3).

### Statistical Analysis

Descriptive statistics were used to characterize the overall study population and the populations from each 2-year cycle, and graphical representations were used to demonstrate temporal trends. The generalized estimation equation (GEE) method was applied to assess the predictors of different outcomes while accounting for the fact that multiple observations might have been available for the same individuals at different cycles. Specifically, an exchangeable within-person working correlation structure was specified in each GEE model and 95% confidence intervals were calculated using a robust variance estimator. Models used to evaluate dichotomous outcomes included a specification for the binomial distribution of the dependent variable, and models used to evaluate continuous outcomes included a specification for the Gaussian distribution of the dependent variable. The independent variables included cycle, sex, race and ethnicity, and age group, and each variable was treated as categorical in the GEE model. To simplify the analysis and avoid multiple comparisons, in the GEE modeling section, we only used the operational hypertension definition that used BP measurement from a single randomly chosen visit during a 2-year cycle. All statistical tests were 2-sided, with a level of significance of 0.05. Data collection using the OMOP model was conducted with the Microsoft SQL Server and data analysis was performed using R (version 4.2.3). The study was approved by the Institutional Review Board at Eastern Virginia Medical School. The study was reported following the STROBE (Strengthening the Reporting of Observational Studies in Epidemiology) reporting guidelines.^11^

## Results

### Population Characteristics

A total of 1,376,325 unique adults met the overall study inclusion criteria. The demographics of the overall study population in each 2-year cycle are shown in Table 1. Each 2-year cycle was analyzed independently, and individual patients could appear in one or more 2-year cycles, depending on the care they received. Thus, the number of patients in the six 2-year cycles varied and ranged from 395,859 patients to 631,892 patients per 2-year cycle. The numbers of outpatient visits and outpatient BP measurements per patient during a 2-year cycle were relatively constant during the study period. The median number of outpatient visits per patient was 2 during 2010-2011 and 3 during the remaining 2-year cycles. The median number of outpatient BP measurements per patient was 5 during 2012-2013 and 2018-2019 and was 4 during the remaining 2-year cycles.

**Table 1.** Demographic characteristics and mean systolic blood pressure (SBP) and mean diastolic blood pressure (DBP) of all patients in each 2-year cycle.

| 2-year Cycle | 2010-2011 | 2012-2013 | 2014-2015 | 2016-2017 | 2018-2019 | 2020-2021 |
| --- | --- | --- | --- | --- | --- | --- |
| Patients | 395,859 | 429,754 | 466,692 | 631,892 | 608,497 | 619,023 |
| Sex |  |  |  |  |  |  |
| Female | 231,203<br>(58%) | 254,610<br>(59%) | 275,386<br>(59%) | 370,352<br>(59%) | 355,759<br>(58%) | 362,140<br>(59%) |
| Male | 164,625<br>(42%) | 175,130<br>(41%) | 191,306<br>(41%) | 261,540<br>(41 %) | 252,736<br>(42%) | 256,874<br>(41%) |
| Race/Ethnicity |  |  |  |  |  |  |
| Non-Hispanic Black | 99,779<br>(25%) | 109,183<br>(25%) | 122,477<br>(26%) | 142,447<br>(23%) | 135,880<br>(22%) | 146,162<br>(24%) |
| Non-Hispanic White | 259,665<br>(66%) | 282,167<br>(66%) | 305, 112<br>(65%) | 438,170<br>(69%) | 423,892<br>(70%) | 423, 186<br>(68%) |
| Hispanic/Latino | 7,342<br>(1.9%) | 11,549<br>(2. 7%) | 13,060<br>(2.8%) | 19,449<br>(3.1%) | 19,691<br>(3.2%) | 21,297<br>(3.4%) |
| Asian | 7,616<br>(1.9%) | 9,294<br>(2.2%) | 10,406<br>(2.2%) | 12,698<br>(2.0%) | 12,834<br>(2.1%) | 13,363<br>(2.2%) |
| Unknown | 21,457<br>(5.4%) | 17,561<br>(4.1%) | 15,637<br>(3.4%) | 19,128<br>(3.0%) | 16,200<br>(2.7%) | 15,015<br>(2.4%) |
| Mean Age (SD) | 51.0 (18.1) | 51.5 (18.2) | 52.3 (18.2) | 52.9 (18.4) | 54.6 (18.2) | 55.3 (18.2) |
| Age Group |  |  |  |  |  |  |
| 18-44 | 142,539<br>(36%) | 151,749<br>(35%) | 158,628<br>(34%) | 210,073<br>(33%) | 182,608<br>(30%) | 181,485<br>(29%) |
| 45-64 | 156,016<br>(39%) | 165,571<br>(39%) | 178, 135<br>(38%) | 234,314<br>(37%) | 224,434<br>(37%) | 221,173<br>(36%) |
| 65-74 | 54,364<br>(14%) | 64,801<br>(15%) | 76,065<br>(16%) | 109,058<br>(17%) | 114,851<br>(19%) | 124,125<br>(20%) |
| >75 | 42,940<br>(11%) | 47,633<br>(11%) | 53,864<br>(12%) | 78,447<br>(12%) | 86,604<br>(14%) | 92,240<br>(15%) |
| Mean SBP (SD) | 125.2 (14.9) | 124.8 (14.8) | 125.1 (14.8) | 126.1 (14.8) | 126.1 (14.2) | 127.5 (14.6) |
| Mean DBP (SD) | 74.7 (9.4) | 74.4 (9.2) | 74.7 (9.1) | 75.5 (9.0) | 75.3 (8.7) | 75.8 (8.8) |

SD=standard deviation

Among all patients, the mean age-adjusted SBP ranged from 125.1 to 127.0 mm Hg and the mean age-adjusted DBP ranged from 74.4 to 76.0 mm Hg (Table 1). Mean SBP and DBP among all patients showed an upward trend during the time of study.

### Hypertension Prevalence and Disparities

The demographics of the hypertensive patients varied slightly depending on the operational hypertension definition. The demographics of hypertensive patients (using the random BP measurement definition, which is comparable to the NHANES study) are shown in eTable 4.

The age-adjusted hypertension prevalence rates over the 12-year study period for each operational hypertension definition are shown in Figure 1. Depending on the operational hypertension definition, the hypertension prevalence rates ranged from 36.5% to 50.9% and prevalence increased by about 5% over the study period regardless of the operational hypertension definition.

**Figure 1.**
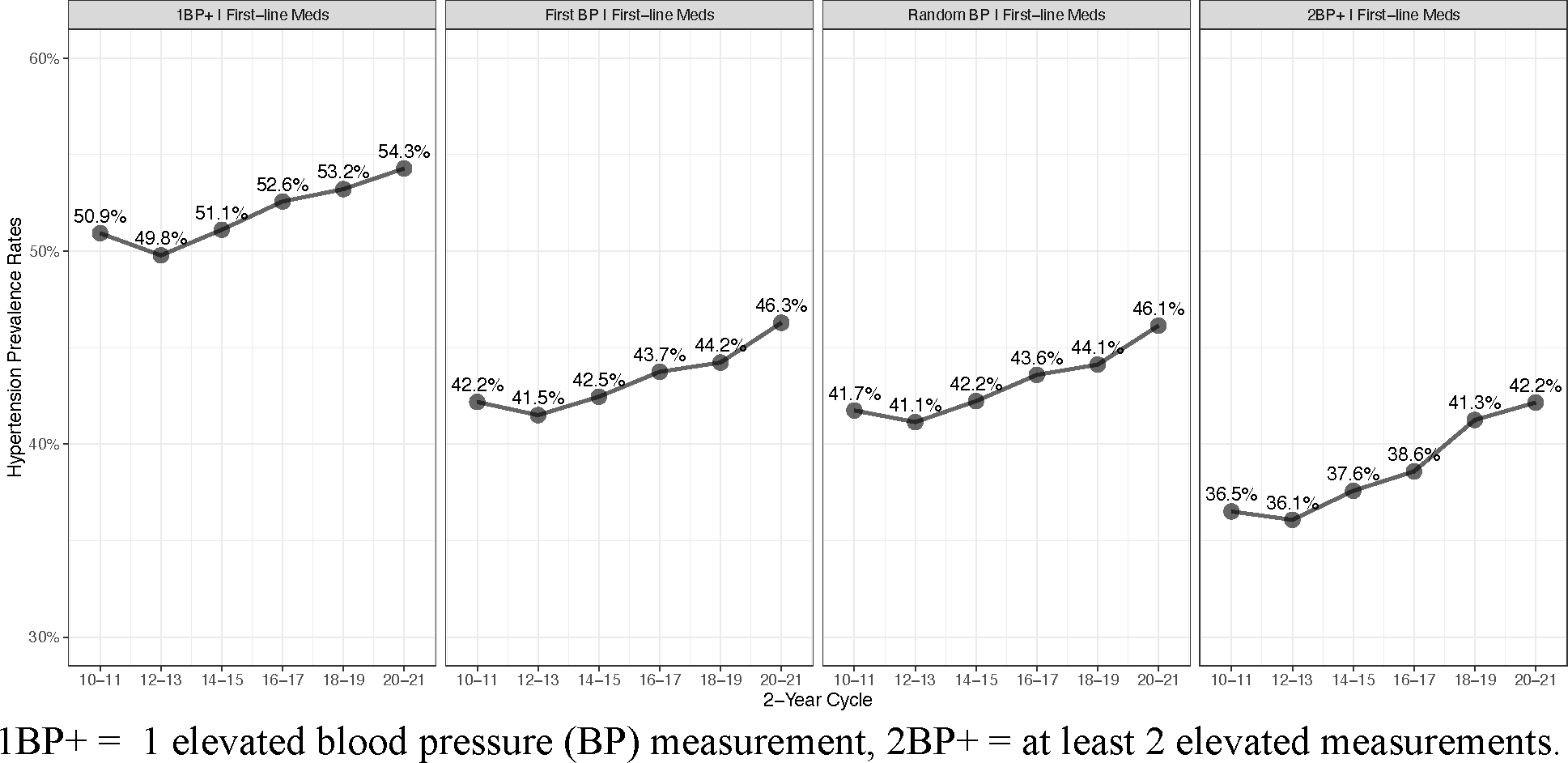
Age-adjusted hypertension prevalence rates in each 2-year cycle by operational hypertension definition.

Non-Hispanic Black patients consistently showed 12-14% higher age-adjusted hypertension prevalence rates compared with the non-Hispanic White patients (OR 2.03, [2.02, 2.04], P <0.001, Figure 2). Hypertension prevalence rates were progressively higher by age group, peaking at 80% in patients >75 years old during 2020-2021 (eFigure 1). Men consistently showed approximately 7% higher hypertension prevalence rates (OR 1.41, [1.40, 1.42], P <0.001, eFigure 2).

**Figure 2.**
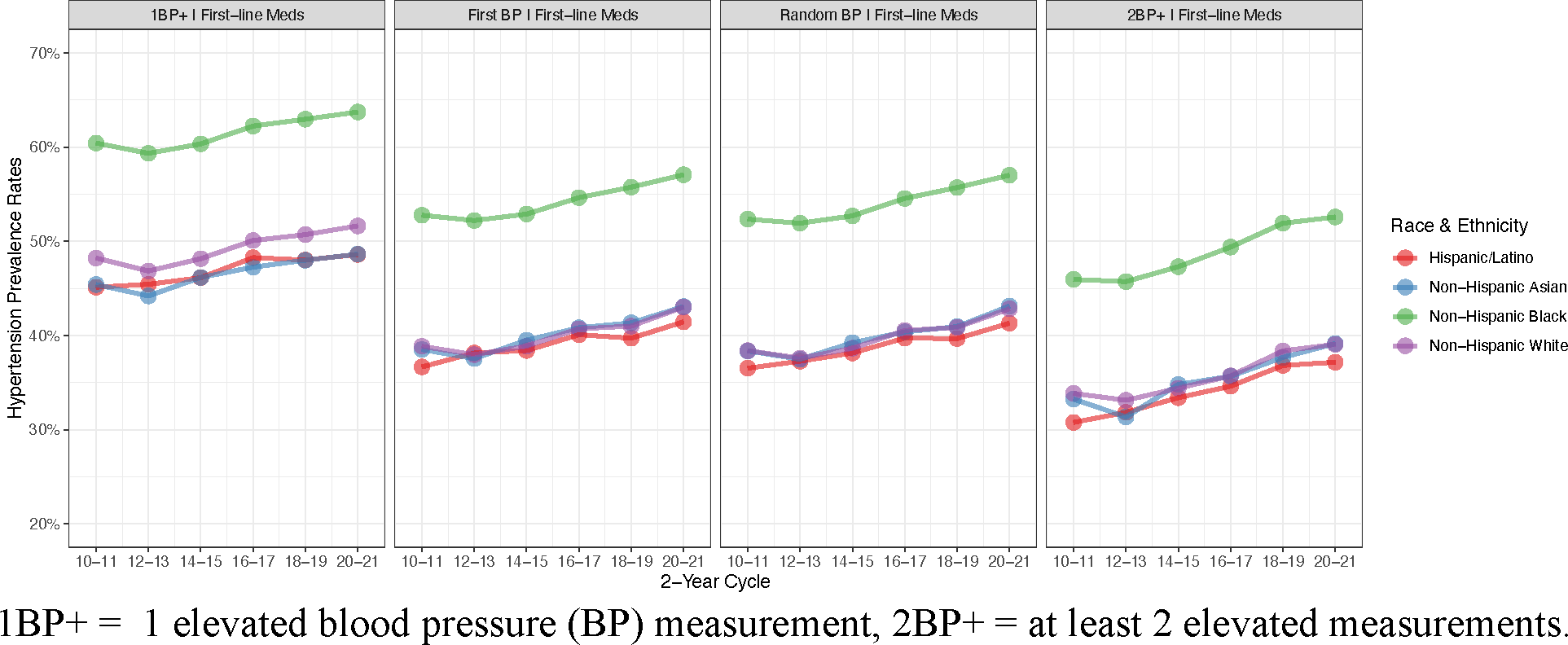
Age-adjusted hypertension prevalence rates by race/ethnicity in each 2-year cycle by operational hypertension definition.

The proportions of hypertensive patients defined by elevated BP only, by use of a first-line antihypertensive medication only, or by both are shown in eFigure3. For all operational hypertension definitions, the proportions of hypertensive patients defined by medication usage or combination of medication usage and elevated BP increased during the study period.

### Blood Pressure Control Rates Among Patients with Hypertension and Disparities

Among patients with hypertension, the age-adjusted BP control rates during the study period ranged from 61.2% to 73.3%, depending on the operational hypertension definition (Figure 3). The trend in BP control rates exhibited an initial increase from 2010-2011 to 2014-2015, followed by a decline in 2016-2017. Thereafter, the control rates rose again from 2018-2019, only to decline once more in 2020-2021. The age-adjusted BP control rates were consistently about 3% lower in men (OR 0.89, [0.88, 0.90], P<0.001, eFigure 4) and consistently about 5-7% lower in non-Hispanic Black patients as compared with non-Hispanic White patients (OR 0.77, [0.76, 0.78], P<0.001, eFigure 5).

**Figure 3.**
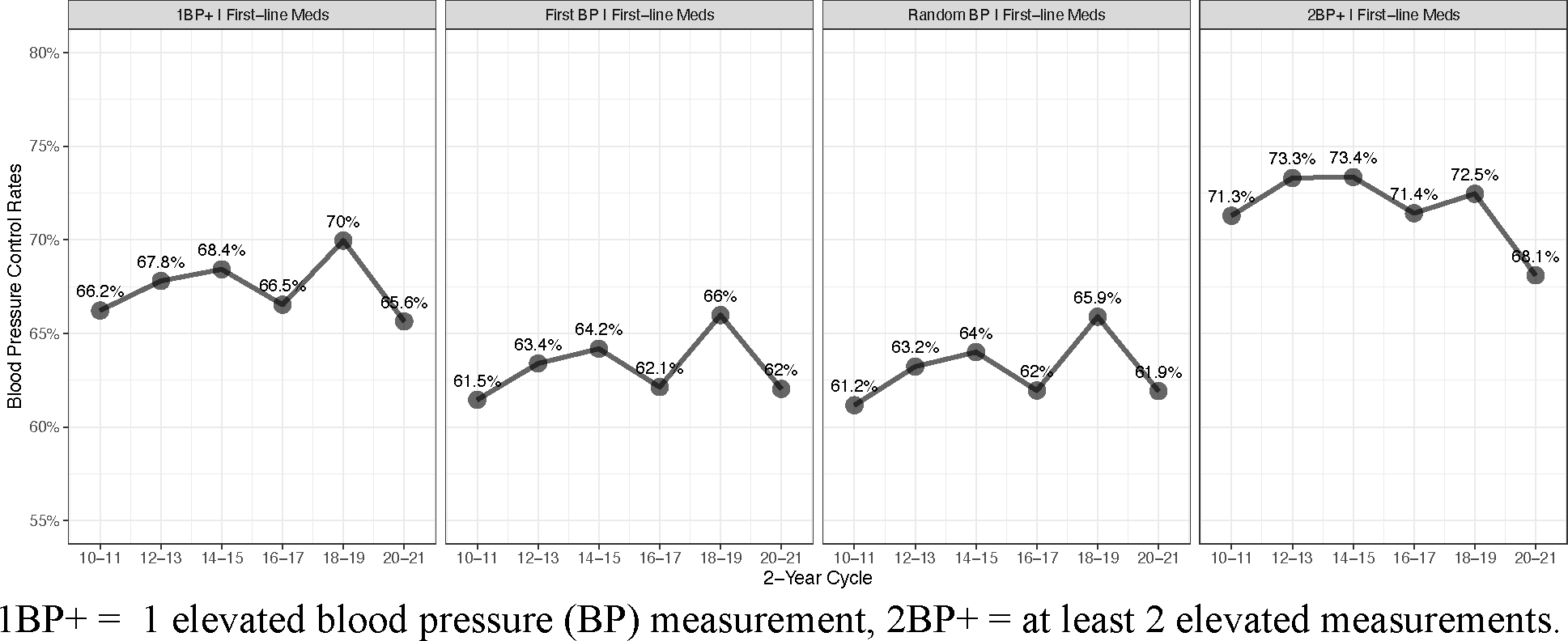
Age-adjusted blood pressure control rates in each 2-year cycle by operational hypertension definition.

**Figure 4.**
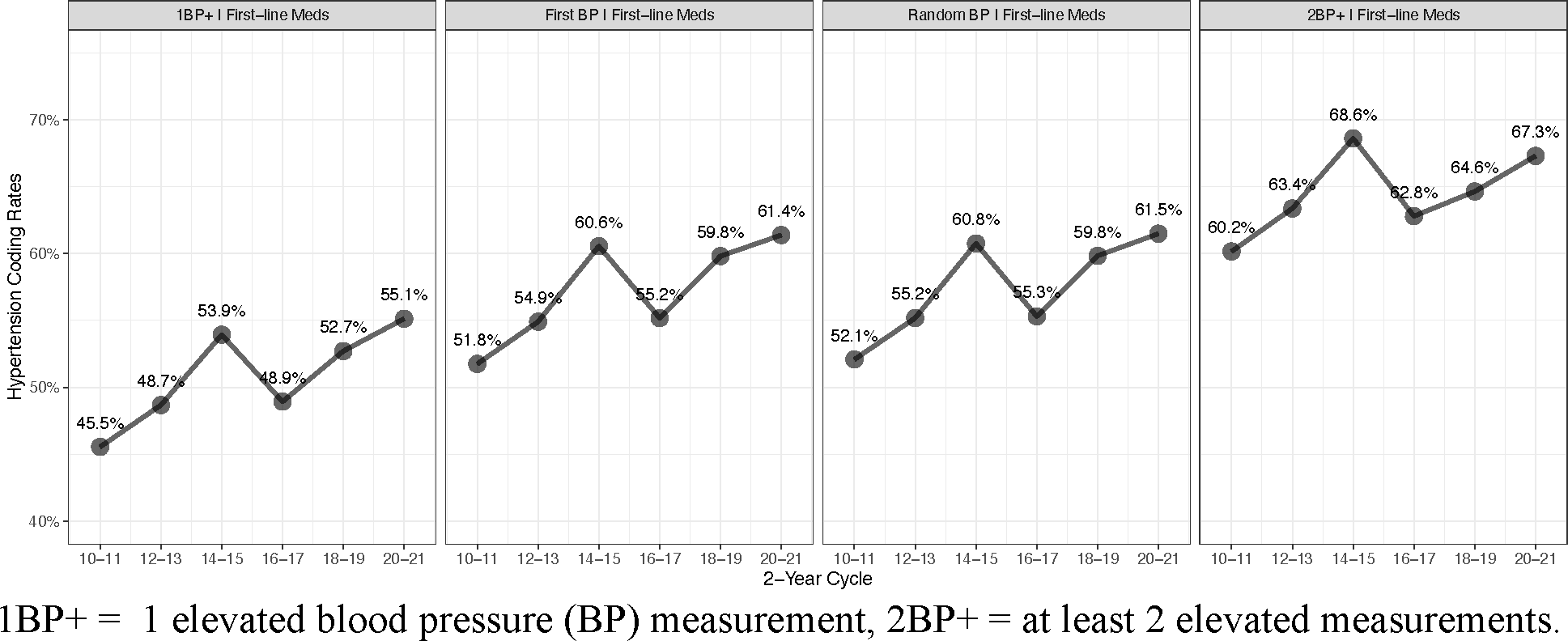
Age-adjusted hypertension coding rates in each 2-year cycle by operational hypertension definition.

Among hypertension patients, the mean age-adjusted SBP ranged from 132.3 to 135.4 mmHg and mean age-adjusted DBP ranged from 76.8 to 78.8 mmHg (eFigure 6). Mean BP was relatively unchanged during the study timeframe regardless of the operational hypertension definition. Age-adjusted mean BP was consistently higher in non-Hispanic Black patients as compared with other non-Hispanic White patients (SBP: +2.61 [2.55, 2.67]; DBP: +1.31 [1.28, 1.35]; P < 0.001 for both, eFigure 7).

### Coded Diagnosis Rates Among Patients with Hypertension and Disparities

Among patients with hypertension, the age-adjusted proportion of patients labelled in the EHR with a coded diagnosis of hypertension ranged from 45.5% to 68.6%, again depending on the operational hypertension definition (Figure 4). The proportion of patients diagnosed with hypertension, as indicated by coded diagnoses, exhibited an initial increase from 2010-2011 to 2014-2015, followed by a decline in 2016-2017. Thereafter, the proportions rose again in 2020-2021. The proportion of patients with a coded diagnosis of hypertension was highest in hypertension groups defined by 2 or more BP measurements than in groups defined by the other definitions. The proportion of patients with hypertension with a coded diagnosis of hypertension was similar in women and men. Rates of receiving a coded diagnosis were consistently about 5% higher in non-Hispanic Black patients (OR 1.26 [1.25, 1.27], P<0.001) and Asian patients (OR 1.31 [1.28, 1.35], P<0.001), compared with White and Hispanic patients (eFigure 8).

## Discussion

In this serial cross-sectional study, we leveraged EHRs of a large health system to analyze regional trends in hypertension using various operational hypertension definitions. We showed a marked increase in age-adjusted hypertension prevalence rates, modest age-adjusted BP control rates, and relatively low but upwardly trending age-adjusted rates of coding hypertension in the EHR during the 12 years of study. In addition, we noted important disparities in age-adjusted hypertension prevalence rates, age-adjusted BP control rates, and age-adjusted mean BP levels that stayed consistent over time. The age-adjusted hypertension prevalence rates were 12-14% higher in non-Hispanic Black patients, 7% higher in men, and progressively higher by age group throughout the study period.

This study demonstrates the ability to observe hypertension trends and disparities at a regional health system level using real-world data, which could greatly facilitate locally designed system-level interventions to address hypertension and racial disparities. We were able to create an agile analytical data platform that enabled a comparison of a variety of operational hypertension definitions and an evaluation of how age, sex, and race/ethnicity affect hypertension trends. Our analytical data platform enabled examination of multiple operational hypertension definitions and we demonstrated substantial differences in outcomes, depending on the sensitivity of the hypertension definition.

Our study demonstrated a steady rise of about 5% in the age-adjusted hypertension prevalence rates throughout the study period, regardless of the operational hypertension definition. Increases in hypertension prevalence rates were seen in all age groups, both sexes, and in all race/ethnicity groups.

The average age of patients and the age adjusted mean SBP and DBP increased during the study period, which may have contributed to the increase in hypertension prevalence rates. The increase in hypertension prevalence rates did not appear to be artifactually affected by the frequency of outpatient visits or BP measurements.

Our definition of hypertension included whether patients were prescribed first-line antihypertension medications, which may have also contributed to the increase in hypertension prevalence rates. The proportion of patients who were defined as hypertensive by the medication-use criterion increased during the 2012-2013, 2018-2019 and 2020-20201 2-year cycles, possibly due to changing guidelines for hypertension. The 2017 ACC/AHA published guidelines recommended a lower BP goal which could have caused more patients to receive first-line antihypertensive medications and could have affected the hypertension prevalence rates during 2018-2019 and 2020-2021.^10^

The age-adjusted proportion of patients with controlled BP rose from 61-71% in 2010-2011 to 64-73% in 2014-2015, fell in 2016-2017, rose again to 66-73% in 2018-2019, and fell in 62-68% in 2020-2021. The pattern was consistent among patients defined by all 4 operational hypertension definitions. Non-Hispanic Blacks had BP control rates that were consistently about 5% lower than Whites. The rise in control rates in all groups during 2018-2019 could be explained by the 2017 ACC/AHA guideline recommendations for tighter BP control.^10^ The fall in control during 2020-2021 may have been caused by changes in healthcare associated with the COVID-19 pandemic.

The proportion of patients with a coded diagnosis of hypertension among patients with hypertension increased during the study period except during 2016-2017 and the trends were similar in all 4 operational hypertension definition groups. There were no differences by sex. Non-Hispanic Blacks and Asians were 6-10% more likely to have a coded diagnosis of hypertension as compared with other race/ethnicity groups. Interestingly, non-Hispanic Black patients were significantly more likely to have hypertension by all the operational hypertension definitions and less likely to have BP controlled, even though they were more likely to have been labelled in the EHR with a coded diagnosis of hypertension.

Our results from a regional health system provide supplemental information to the national estimates reported by the National Health and Nutrition Examination Survey (NHANES). Using a comparable operational hypertension definition (a single random visit BP measurement), our age-adjusted hypertension prevalence rates are higher (41.1% to 46.1%) and increased steadily as compared with NHANES which reported prevalence rates that were lower and remained constant (30% to 32%).^2^ Our BP control rates (61.2% to 65.9%) were also higher than those reported by NHANES (31.8% to 53.8%). These differences may be because our study population was derived from patients exposed to a healthcare system rather than subjects randomly selected from the general population. Importantly, the proportion of non-Hispanic Blacks in our study was roughly twice that from the NHANES study, reflecting the demographics of our region, which may have increased the hypertension prevalence rates.

Studies have suggested that hypertension prevalence based on methods using electronic health records could be underestimated.^12^ In our study, we used the EHR, but we defined hypertension based on measurement of BP, rather than based on the coded diagnosis of hypertension, which appears to have overcome the potential of under-reporting hypertension using the EHR.

Our hypertension control rates were similar to those reported by the PCORnet Blood Pressure Control Laboratory, which reported an average BP control rate of 62%.^13^ Their findings from 25 health systems showed different demographics than our regional analysis, with Black patients only comprising 15% of their population, as compared to approximately 30% of the hypertensive patients in our study. While nationwide studies are informative, limiting the analysis to a single regional health system may provide actionable insights regarding disparities that are more relevant locally, based on the demographics of that regional health system.

Banerjee et al measured the hypertension coding rate in 251,590 patients defined hypertension by 2 or more BP readings ≥140/90 mm Hg and/or antihypertensive medications prescribed during the study period. Among those patients, the coded diagnosis rate was 62.9%, very similar to our finding among patients with a comparable operational hypertension definition.^12^

Our study has recognized strengths and limitations. Our approach of analyzing real-world data from a single health system is practical and feasible, and the results are similar to the multicenter approach of the PCORnet Blood Pressure Control Laboratory.^14^ Our data platform allows rapid evaluation of populations at scale and could be a source of continuing evaluation with planned periodic additions to the data base,^15,16^ providing a powerful tool for generating actionable insights from real-world data.^17,18^ This data resource is an model of what could be a useful generator of insights for a learning health system.^19,20^

A possible limitation of our study, as compared with the NHANES approach,^2^ however, is that BP was obtained in the usual care setting, rather than through a rigorous protocolized approach. Although clinicians expect and often demand accurate BP measurements in usual care settings, the BP measurements in real-world settings may be less accurate than in the protocolized setting of a prospective research study.^21^ Also, the coded diagnosis of hypertension in the EHR could have been affected by recall bias and other sources of possible coding error.^15^

In conclusion, we were able to use real-world EHR data to establish hypertension prevalence rates, control rates, and diagnostic coding rates in a large population using the EHR from a large regional health system, showing important trends and opportunities to address racial disparities. Our study may provide a model for other health systems seeking to improve hypertension care at a regional health system level and may lead to further research on how to use real-world data and computational approaches for addressing hypertension.

## Data Availability

The primary data is protected health information and cannot be made available.

