## Supplemental Material for "Hypertension Trends and Disparities over Twelve Years in a Large Health System: Leveraging the Electronic Health Records"

### On-Line Supplemental Material

**eTable 1.** Drug names and OMOP concept identification numbers for first-line antihypertension drug classes.

| Class | Drug Name (Concept ID) |
|---|---|
| Thiazide-like diuretics | Chlorthalidone (1395058); HCTZ (974166); Indapamide (978555); Metolazone (907013) |
| Angiotensin converting enzyme inhibitors | Benazepril (1335471); Captopril (1340128); Enalapril (1341927); Fosinopril (1363749); Lisinopril (1308216); Moexipril (1310756); Perindopril (1373225); Quinapril (1331235); Ramipril (1334456); Trandolapril (1342439) |
| Angiotensin receptor blockers | Azilsartan (40235485); Candesartan (1351557); Eprosartan (1346686); Irbesartan (1347384); Losartan (1367500); Olmesartan (40226742); Telmisartan (1317640); Valsartan (1308842) |
| Dihydropyridine calcium channel blockers | Amlodipine (1332418); Felodipine (1353776); Isradipine (1326012); Nicardipine (1318137); Nifedipine (1318853); Nisoldipine (1319880) |
| Non-dihydropyridine calcium channel blockers | Diltiazem (1328165); Verapamil (1307863) |

**eTable 2.** Proportions of patients in each age group used for age-adjustment in the overall population and the hypertensive patients by operational hypertension definition.

| | Overall Population | Operational Hypertension Definition | | | |
|---|---|---|---|---|---|
| Age Group | | Any 1 BP elevation | First BP measurement | Random BP measurement | At least 2 BP elevation |
| 18-44 | 33.0 | 16.7 | 14.7 | 14.6 | 12.1 |
| 45-64 | 37.6 | 41.9 | 42.2 | 42.1 | 42.4 |
| 65-74 | 16.9 | 22.9 | 23.7 | 23.7 | 24.8 |
| >75 | 12.5 | 18.5 | 19.5 | 19.5 | 20.7 |

**eTable 3.** Hypertension coding terms and corresponding OMOP concept identification numbers.

| Term | Concept ID |
|---|---|
| Essential hypertension | 320128 |
| Hypertension secondary to endocrine disorder | 4110948 |
| Secondary hypertension | 319826 |
| Renovascular hypertension | 317895 |
| Hypertensive urgency | 40481896 |
| Hypertensive emergency | 43020424 |
| Renal hypertension | 443771 |
| Pre-existing hypertension in obstetric context | 4311246 |
| Hypertensive crisis | 45768449 |
| HELLP syndrome | 4316372 |

| | |
|---|---|
| Malignant essential hypertension | 317898] |

**eTable 4.** Demographic characteristics of patients with hypertension defined by a single random visit blood pressure measurement in each 2-year cycle.

| 2-year Cycle | 2010-2011 | 2012-2013 | 2014-2015 | 2016-2017 | 2018-2019 | 2020-2021 |
|---|---|---|---|---|---|---|
| Patients | 159,138 | 171,875 | 194,474 | 275,124 | 277,555 | 297,845 |
| Sex | | | | | | |
| Female | 85,714 (53.9%) | 93,283 (54.3%) | 105,103 (54.0%) | 147,258 (53.5%) | 148,541 (53.5%) | 159,023 (53.4%) |
| Male | 73,412 (46.1%) | 78,586 (45.7%) | 89,371 (46.0%) | 127,866 (46.5%) | 129,014 (46.5%) | 138,818 (46.6%) |
| Race/Ethnicity | | | | | | |
| Non-Hispanic Black | 48,974 (30.8%) | 53,317 (31.0%) | 61,490 (31.6%) | 74,259 (27.0%) | 74,770 (26.9%) | 82,560 (27.7%) |
| Non-Hispanic White | 98,647 (62.0%) | 106,470 (61.9%) | 120,514 (62.0%) | 183,994 (66.9%) | 186,142 (67.1%) | 197,085 (66.2%) |
| Hispanic/Latino | 2,064 (1.3%) | 3,187 (1.9%) | 3,782 (1.9%) | 5,920 (2.2%) | 6,242 (2.2%) | 7,259 (2.4%) |
| Asian | 2,672 (1.7%) | 3,301 (1.9%) | 3,897 (2.0%) | 4,883 (1.8%) | 5,143 (1.9%) | 5,754 (1.9%) |
| Other/Unknown | 6,781 (4.3%) | 5,600 (3.3%) | 4,791 (2.5%) | 6,068 (2.2%) | 5,258 (1.9%) | 5,187 (1.7%) |
| Mean Age | 59.1 (15.6) | 59.9 (15.4) | 60.5 (15.3) | 61.0 (15.4) | 62.4 (15.0) | 62.7 (15.0) |
| Age Group | | | | | | |
| 18-44 | 26,995 (17.0%) | 27,122 (15.8%) | 28,883 (14.9%) | 40,596 (14.8%) | 34,906 (12.6%) | 38,091 (12.8%) |
| 45-64 | 72,114 (45.3%) | 76,123 (44.3%) | 84,206 (43.3%) | 113,853 (41.4%) | 111,293 (40.1%) | 114,432 (38.4%) |
| 65-74 | 32,103 (20.2%) | 37,465 (21.8%) | 45,238 (23.3%) | 67,081 (24.4%) | 71,558 (25.8%) | 79,830 (26.8%) |
| >75 | 27,927 (17.5%) | 31,165 (18.1%) | 36,147 (18.6%) | 53,594 (19.5%) | 59,798 (21.5%) | 65,492 (22.0%) |

**eFigure 1.** Age-adjusted hypertension prevalence rates by age groups in each 2-year cycle by operational hypertension definition.

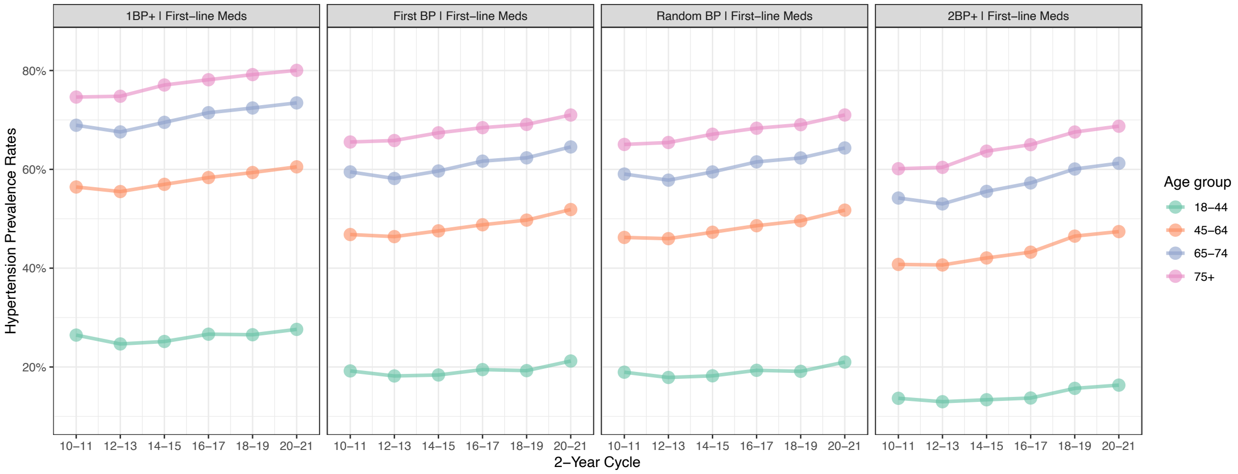


1BP+ = 1 elevated blood pressure (BP) measurement, 2BP+ = at least 2 elevated measurements.

**eFigure 2**. Age-adjusted hypertension prevalence rates by sex in each 2-year by operational hypertension definition.

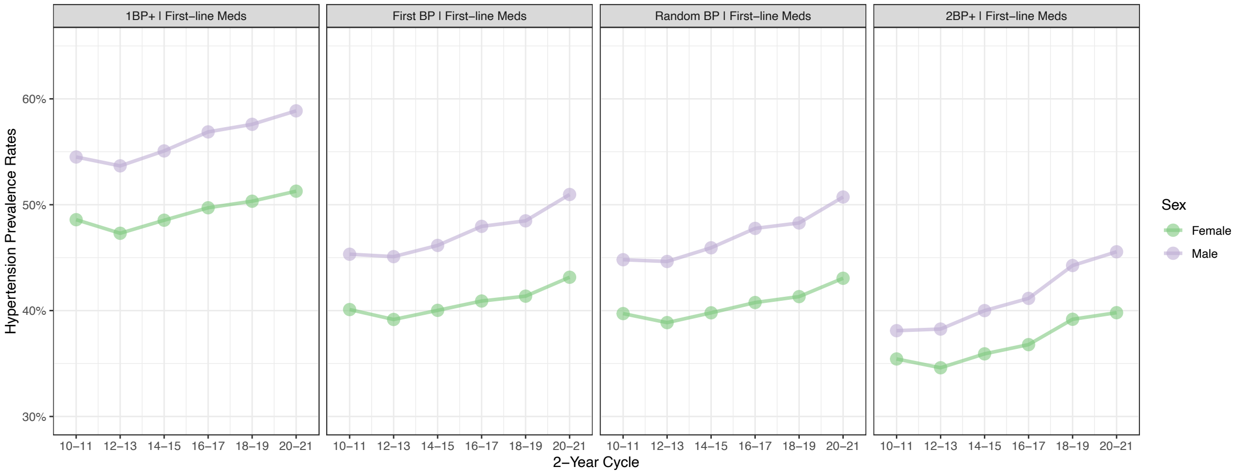


1BP+ = 1 elevated blood pressure (BP) measurement, 2BP+ = at least 2 elevated measurements.

**eFigure 3.** Composition of the components of each operational hypertension definition in each 2-year cycle.

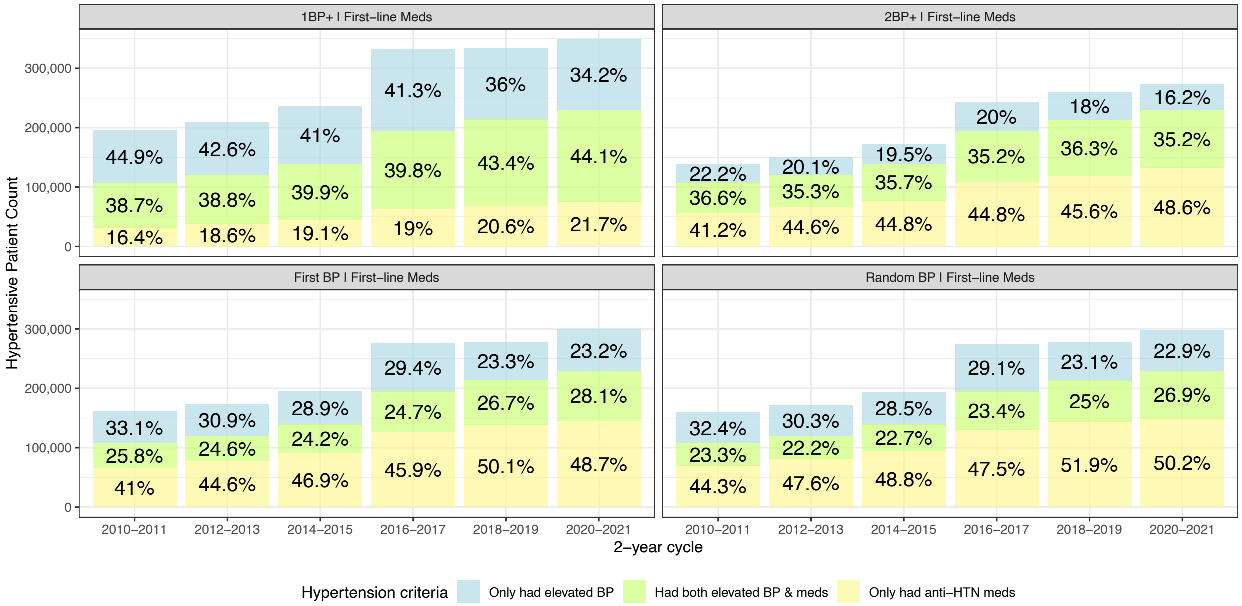


**eFigure 4.** Age-adjusted blood pressure control rates by sex in each 2-year cycle by operational hypertension definition.

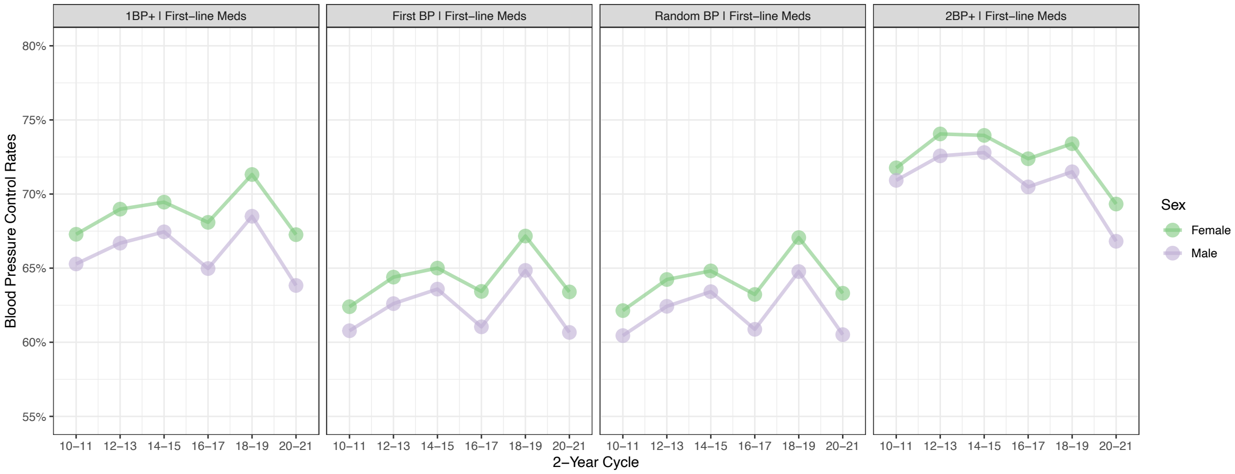


1BP+ = 1 elevated blood pressure (BP) measurement, 2BP+ = at least 2 elevated measurements.

**eFigure 5.** Age-adjusted blood pressure control rates by race/ethnicity in each 2-year cycle by operational hypertension definition.

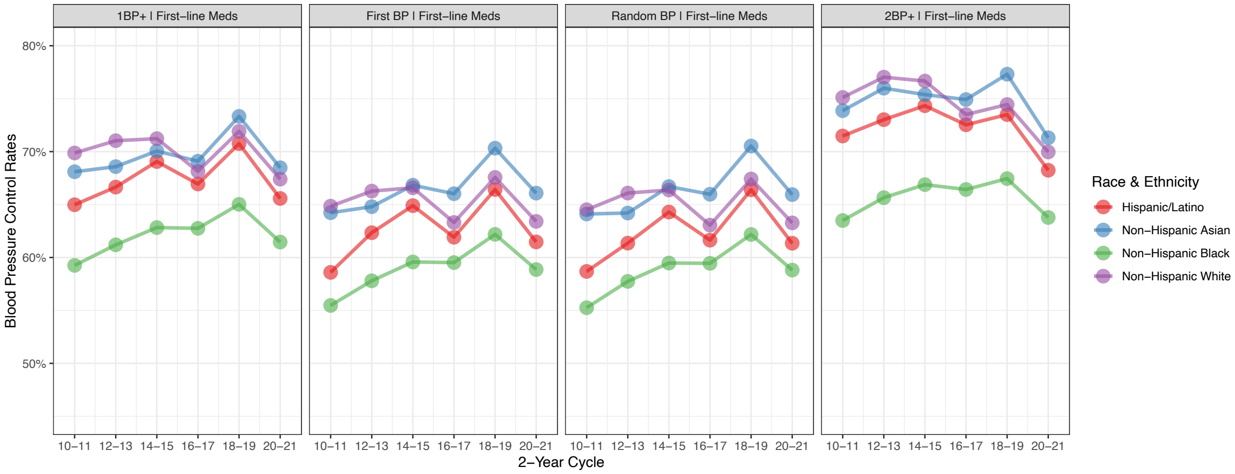


1BP+ = 1 elevated blood pressure (BP) measurement, 2BP+ = at least 2 elevated measurements.

**eFigure 6.** Age-adjusted mean systolic and diastolic blood pressures in hypertensive patients in each 2-year cycle by operational hypertension definition.

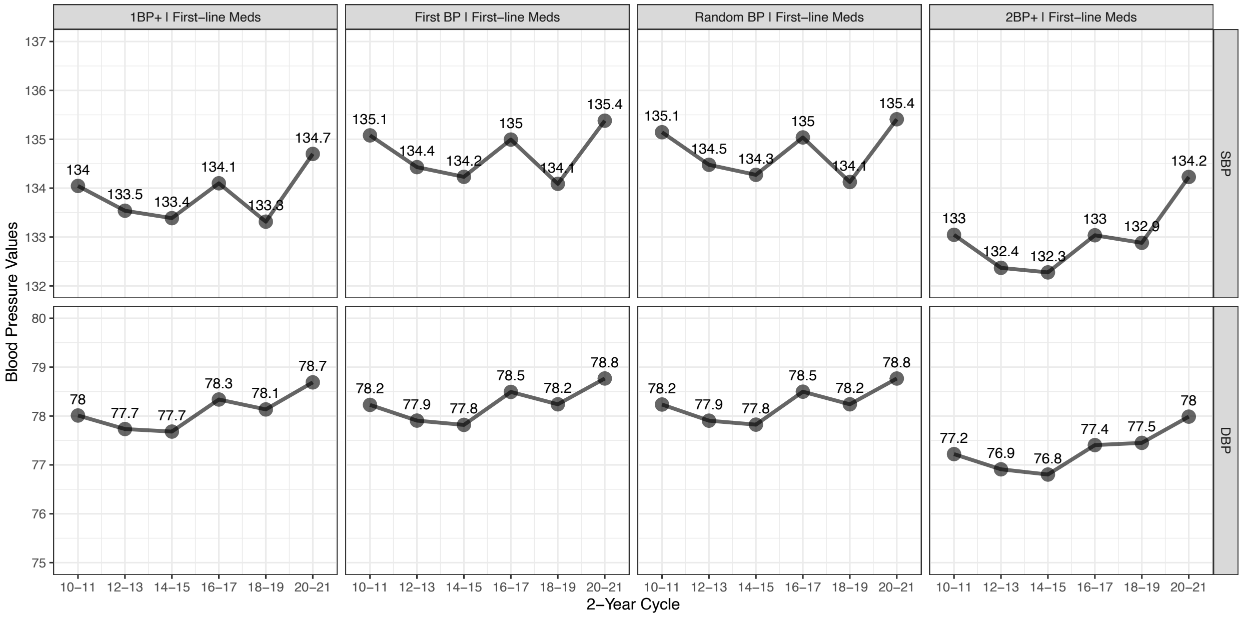


1BP+ = 1 elevated blood pressure (BP) measurement, 2BP+ = at least 2 elevated measurements,
SBP = systolic blood pressure, DBP = diastolic blood pressure.

**eFigure 7.** Age-adjusted mean systolic and diastolic blood pressure by race/ethnicity in hypertensive patients in each 2-year cycle by operational hypertension definition.

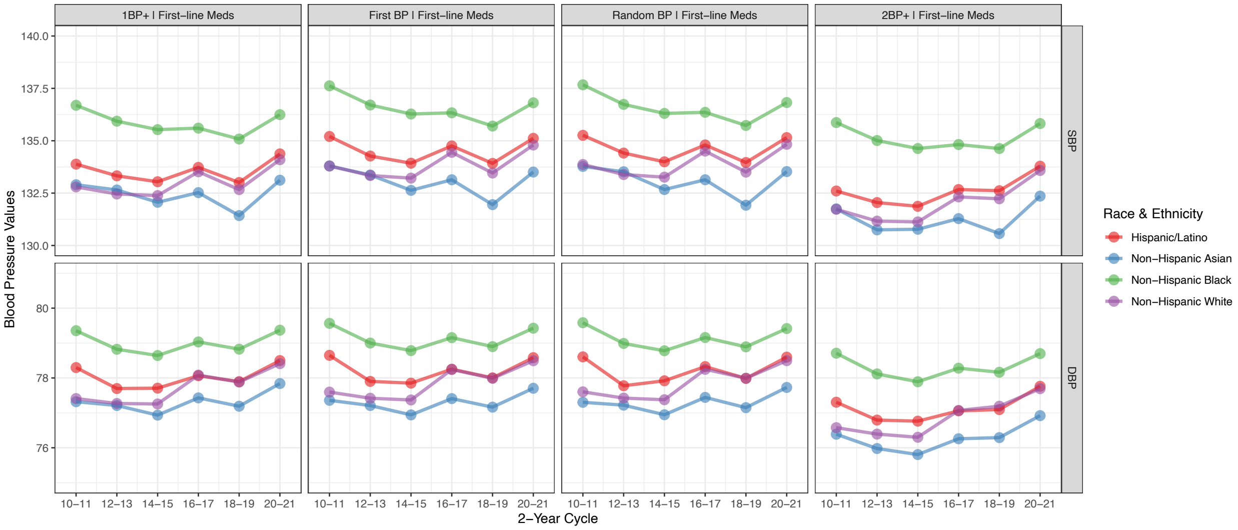


1BP+ = 1 elevated blood pressure (BP) measurement, 2BP+ = at least 2 elevated measurements, SBP = systolic blood pressure, DBP = diastolic blood pressure.

**eFigure 8.** Age-adjusted hypertension coding rates by race/ethnicity in each 2-year cycle by operational hypertension definition.

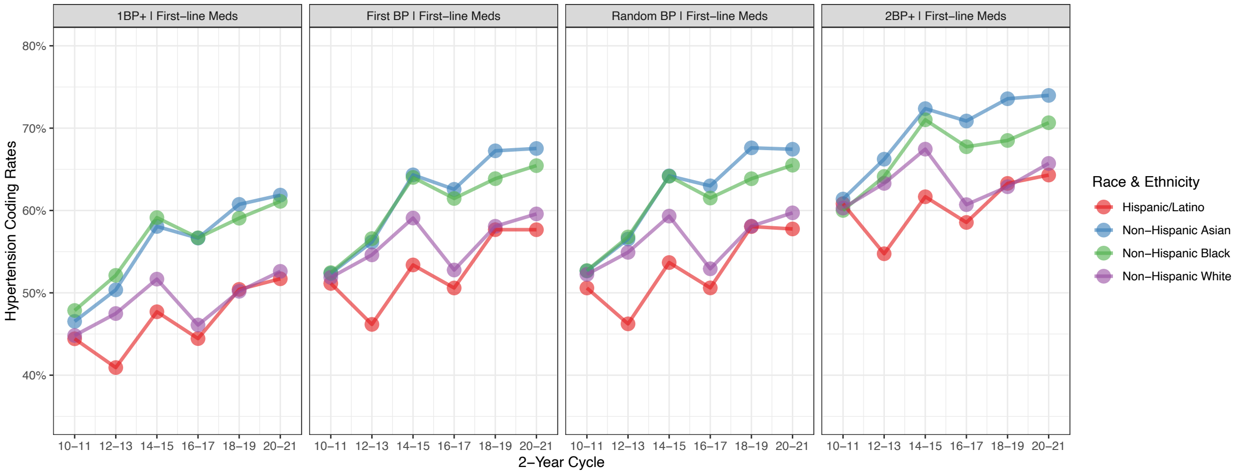


1BP+ = 1 elevated blood pressure (BP) measurement, 2BP+ = at least 2 elevated measurements.
